# A single-blinded randomized crossover trial comparing peer-to-peer and standard instruction on airway management skill training

**DOI:** 10.1101/19001768

**Authors:** Usapan Surabenjawong, Paul Edward Phrampus, John Lutz, Deborah Farkas, Apoorva Gopalakrishna, Apichaya Monsomboon, Chok Limsuwat, John Marc O’Donnell

## Abstract

**Background:** Peer-to-peer teaching, which is an alternative to standard teaching (by expert instructors), has the potential to emphasize student self-learning and reduce the cost and workload of the instructor. Self-instruction videos with peer feedback are highlighted in many medical and nursing school curricula.

**Objective:** To evaluate whether peer to peer instruction supported by a structured curriculum and video exemplars is not inferior to standard instructor-led teaching in basic airway management skill, knowledge, and confidence attainment.

**Method:** This single blinded randomized crossover trial was conducted with a sample of novice nursing students. Data was collected through the pre-to post-knowledge and confidence assessments. The students were randomly assigned to two crossover groups. Each student learned basic airway management skills through both methods. The students’ performances were recorded in every session with recordings reviewed by blinded expert instructors.

**Results:** The study included 48 participants, who were assigned into both the expert instruction group and peer-to-peer group through computer generated randomization. The skill rating scores of the peer-to-peer group were not inferior to the standard teaching. With further analysis, we noted that the peer-to-peer group scores had significantly higher scores demonstrating a large effect size (Cohen’s *d* of 1.07 (p-value 0.002) for oropharyngeal airway insertion, 1.14 (p-value <0.001) for nasopharyngeal airway insertion and 0.81 (p-value 0.003) for bag mask ventilation). There was no significant difference between pre- and post-knowledge scores across groups (p-value of 0.13 and 0.22 respectively). Participants in both groups reported higher confidence after learning. However, the difference was not statistically significant.

**Conclusions:** Undergraduate nursing students trained in basic airway management skills by peer-to-peer instruction and a structured curriculum did not show inferior scores compared to the students who were trained by expert instructors. There was no significant difference in the knowledge and confidence levels between the groups.

## INTRODUCTION

Basic airway management is an essential skill for a wide variety of healthcare providers. Nowadays, these skills are commonly taught by expert instructors who lead face-to-face teaching sessions.^1^ There are several disadvantages to this approach, including the cost of obtaining qualified instructors. Expert instructors may also prolong sessions by providing didactic information, thus limiting actual practice time. Another limitation of training by expert is logistical in nature due to the need to schedule training around the often fixed schedules of these subject matter experts. Structured curricula, including the use of self-instruction videos, has been shown to be as effective as standard face-to-face training in certain circumstances such as for teaching basic airway management, cervical collar application, emergency extremity splinting skills, manual cardiac defibrillation, and automated external defibrillator.^2,3^ In these situations, trainees have demonstrated comparable confidence and psychomotor learning outcomes when compared to a traditional teaching approach. Peer-to-peer instruction is also an approach that has been demonstrated to be effective. Concepts of self-reflection, self-assessment, peer-assessment and peer-feedback are currently thought to be the principles that underlie this approach across multiple applications for medical curricula.^4^ Teaching students the process of reflective practice through peer-to-peer interaction is thought to stimulate critical thinking, assist with problem-solving and helps to emphasize the lifelong skill of self-directed learning. The peer-to-peer approach also helps participants gain new understanding, offers a fresh perspective and reveals alternative training pathways that may help to accelerate future skill acquistion.^5^

There have been no reports in the literature of a structured curriculum model for peer-to-peer instruction of basic airway management for nursing students although other skills ranging from obtaining a blood pressure to performing sterile technique using a ‘bundled’ curriculum have been reported.^6^ It is hypothesized that a structured curriculum which includes a combination of self-instruction videos, peer-to-peer instruction and guided peer-to-peer feedback using structured debriefing, such as the Gather, Analyze, Summarize or GAS model, will be as effective as standard instructor-led teaching of novice nursing student acquisition of basic airway management knowledge, skill and confidence. The peer-to-peer teaching method has the potential to be a cost-saving alternative for teaching basic airway management. It can help to address the problem of maintaining high quality training in the setting of rapidly increasing number of healthcare student trainees yet a fixed number of instructors. If demonstrated to be effective, this method could be applied for a wide variety of learning objectives across a range of skills or techniques.

The purpose of the study is to evaluate whether peer-to-peer instruction supported by a structured curriculum and video exemplars is not inferior to standard expert instruction of basic airway management in the areas of knowledge, skill and confidence levels.

## METHODS

A single-blinded, randomized crossover trial was conducted with 48 undergraduate nursing students who were between 18 and 65 years old and identified as novice-level with respect to basic airway management. After obtaining exempt IRB approval for the study through the University of Pittsburgh, investigators began recruitment. Subjects were recruited through a broadcast email sent to nursing professors, as well as through recruitment scripts that were placed in public areas at the University of Pittsburgh School of Nursing. After expressing interest, participants contacted the PI, were read a research script and instructed to register for an account through the Peter M. Winter Institute for Simulation, Education and Research (WISER) simulation information management system (SIMS) web-portal. After registration, participants entered the password-protected research portal and completed an online pre-course survey (to attain demographic and attitude data) and a 10-item knowledge quiz. After the pre-course survey and quiz was completed, all participants were directed to review an online self-study module on basic airway management. Completion of this module was verified through the SIMS course-reporting dashboard function. They were then scheduled for learning sessions at WISER. Participants were randomized into two groups (peer-to-peer and expert instructor) through a computer-generated program (Figure 1). The learning sessions were two hours in duration after which the participants were instructed to access the online research course portal and complete the post-course attitude and knowledge evaluations. After completion of the instruments, expert instructors offered a supplemental instructional session to the students who had received peer-to-peer instruction for either bag-valve-mask ventilation or airway management. This was done to ensure that all students had an opportunity to receive ‘standard’ instruction in these skills.

**Figure 1.**
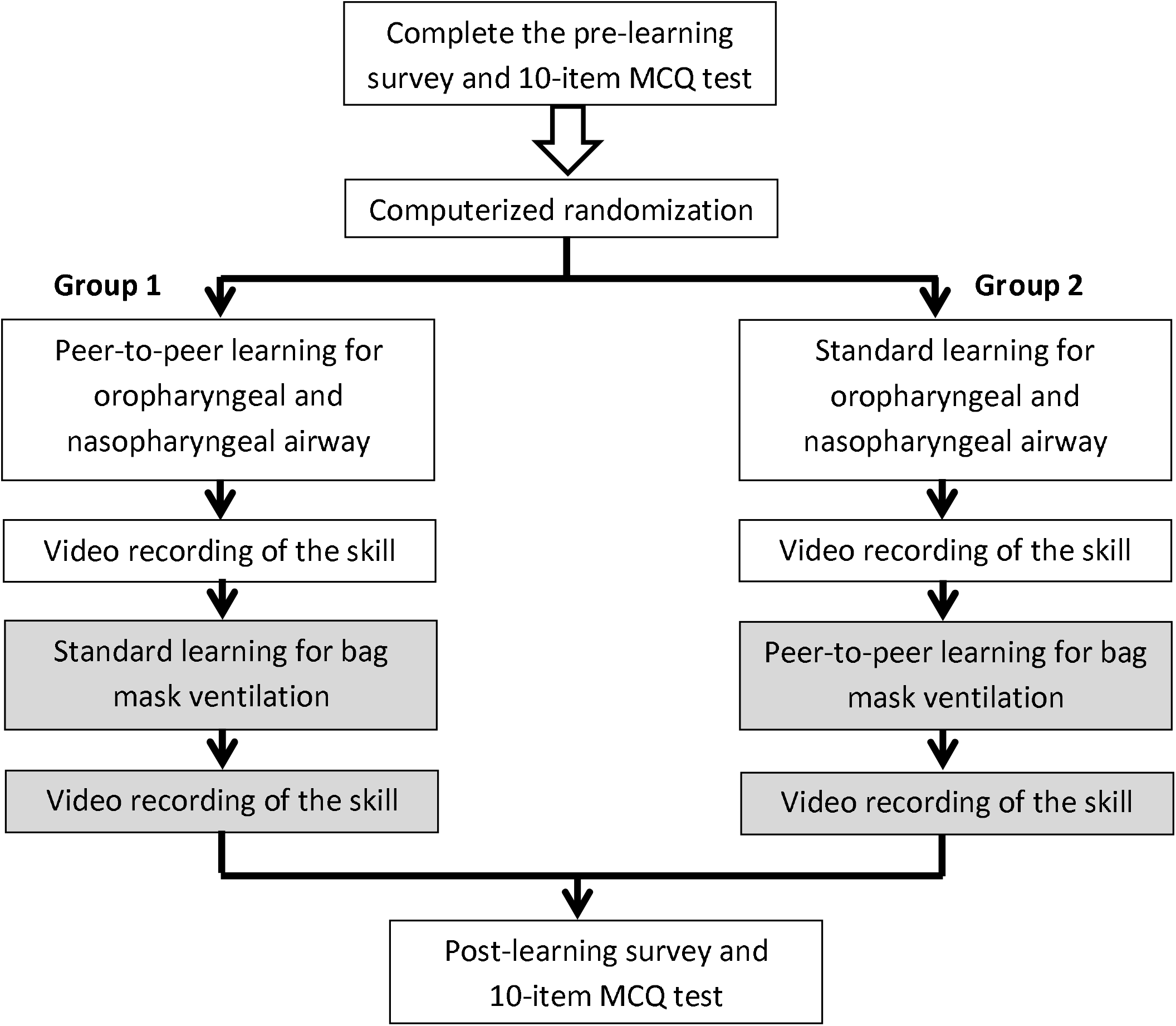
Flow of the study.

### Survey and knowledge test development

The pre- and post-learning surveys were created in a mirror pattern in order to measure four learning components. The four components are: knowledge confidence, stress during class, self-perception of knowledge, and pre-to post-knowledge acquisition. The pre- and post-learning surveys measured the first three components using a Likert scale, in which each item was rated from 1 (strongly disagree) to 5 (strongly agree). In the post-learning survey there was an additional question included which evaluated participants’ preferences between peer-to-peer and standard expert training of basic airway management skills.

Knowledge acquisition was measured with an online 10-item multiple choice question (MCQ) assessment. The content was validated in an iterative fashion by referencing each item to published literature and through expert consensus by three airway experts who are registered nurses and Certified Registered Nurse Anesthetists (CRNAs). The three airway experts were female faculty members of a large US University with an average of 14.7 years of clinical experience (range 8-25 years). These faculty experts were not a part of the research team. After initial validation, the assessment had a test-retest reliability coefficient of 0.82 from pilot tests administered to a separate group of 28 nursing students. No changes were made to assessment items based on the reliability evaluation.

### Development of the skill checklists

The checklists for each skill were developed by using the standard procedural manual from pre-hospital trauma life support as a guideline.^7^ An online Modified Delphi process was used to validate the content and the steps of the checklist. There were 13 airway experts, from three nations with more than 240 years of combined airway experience (mean 20.3, SD 11.7) involved in the modified Delphi process. Each expert reported managing at least 700 airways within the past two years. Overall agreement with the items on the oral airway, nasal airway and bag mask ventilation skills was high ranging from 0.82 to 0.98 average agreement (Table 1). Two blinded independent, attending emergency room physicians working in a large, urban emergency room setting served as the raters for the skill videos. These attending emergency physicians reported airway management experience of greater than 8 years. The inter-rater reliability (by Cohen kappa coefficients) among the raters was 0.77 for oropharyngeal airway, 0.84 for nasopharyngeal airway and 0.81 for bag mask ventilation.

**Table 1.** Overall rater agreement on the skill checklist for the oropharyngeal airway (6 items), nasopharyngeal airway (6 items) and bag mask ventilation (7 items)

| Agreement | Oropharyngeal airway | Nasopharyngeal airway | Bag mask ventilation |
| --- | --- | --- | --- |
| Item 1 | 0.92 | 0.92 | 0.92 |
| Item 2 | 0.92 | 0.92 | 0.92 |
| Item 3 | 0.75 | 0.58 | 1 |
| Item 4 | 0.58 | 1 | 1 |
| Item 5 | 0.83 | 1 | 1 |
| Item 6 | 0.92 | 1 | 1 |
| Item 7 | - | - | 1 |
| **Mean** | **0.82** | **0.90** | **0.98** |

### Detailed Description of Learning Sessions

### Simulation Setting and Devices

All simulation sessions were conducted at the WISER Institute of the University of Pittsburgh. WISER is an 18,000 square foot, state of the art, interprofessional simulation center which hosts more than 15,000 simulation encounters annually. The mannequins used in the study were Laerdal SimMan® (Laerdal Inc., Stavangar Norway) patient simulators because the airway structure realism is similar-enough to a patient for effective training of basic airway techniques. The following airway scenarios were introduced during the various learning sessions:

1. A 32 years old male is brought to your department after a motorcycle accident. He is unconscious and breathing loudly. You want to assist his breathing by inserting an oropharyngeal airway. Please perform this procedure step by step.
2. A 45 years old woman has taken an overdose of sleeping pills and is brought to the emergency room by her husband. She is semi-conscious and snoring. You want to assist her breathing by inserting a nasopharyngeal airway. Please perform this procedure step by step.
3. A 63 years old man has difficulty breathing for 2 days. His diagnosis is severe pneumonia. Your attending staff orders you to give him one person bag mask ventilation. Please perform step by step bag mask ventilation and administer at least 3 breaths

### Study Flow (Figure 1)

Subjects participated in two learning sessions: the oropharyngeal/ nasopharyngeal airway insertion session and the bag-mask ventilation session. Each session was one hour long. Participants were randomly assigned to either the peer-to-peer learning group or the expert learning group. There were 2 students in each group. Half of the participants were assigned to learn the airway (oral and nasal) insertion by peer-to-peer instruction first and then crossed over to the expert instructor teaching group for bag-mask ventilation skills. The first groups were labeled as “Group 1.” The other half of participants learned airway insertion through expert instruction, then crossed over to the peer-to-peer learning group for bag-mask ventilation skill training. This groups were labeled as “Group 2.” After each training session, each participant’s performance was recorded on video without showing the participant’s face or other identifying information. As noted, these videos were later viewed and rated by the 2 blinded observers.

1. *The standard instructor-led teaching method*: The instructors in the standard expert teaching group were four experienced critical care nurses in the third year of a graduate-level nurse anesthesia training program. Each instructor had verified expertise in basic airway management (at minimum 600 successful airway management experiences within 2 years.) All instructors were oriented to teach the basic airway management skills in this study for standardization of teaching techniques prior to the learning sessions. The instructors debriefed each skill attempt using the Gather, Analyze, Summarize or GAS debriefing model which is the recommended model at WISER. The participants in the instructor-led teaching session had time to practice skills as much as they needed under instructor guidance in the allotted one hour time period.
2. *The peer-to-peer learning method*: The peer-to-peer learning method allowed one participant to learn with another participant with the guidance of self-instruction videos. The self-instruction videos contained steps of the specific basic airway management skill, as well as a review of the GAS debriefing model to guide peer review of airway skills. The knowledge content of the self-instruction videos were reviewed for validity by the three Nurse Anesthesia faculty airway experts as well as two experienced simulation educators with more than 30 years of combined experience in simulation education and airway management. After the participants watched the videos, they received a checklist for each basic airway management skill to use as a guide for practice and reflection. The participants in the peer-to-peer learning group also had opportunity to practice the skills and debrief in the allotted one hour.

### Statistical analysis

The standard deviation for basic airway management OSCE scores after use of a self-instruction video has been reported to be 12.41%.^2^ This finding was used to estimate the sample size necessary for the study. On an *a priore* basis, non-inferiority was determined if the lower bound of the 2-sided 95% confidence interval of the mean difference did not exceed a 10% margin. With 80% power (1-β), 0.05 significance level (α) and 10% non-inferiority limit (d), the calculated sample size was 20 students per group.

Categorical data were analyzed using the Fisher exact test. The Wilcoxon signed-rank test and the Mann–Whitney U tests were used for calculation of comparisons between pre- and post-confidence and Likert scale data. Cohen’s kappa coefficient was used to demonstrate the inter-rater reliability for skill assessment. After non-inferiority was established, skill improvement effect sizes between peer-to-peer and expert instruction were calculated using Cohen’s *d*, in which the values of ≥0.80 are considered a large effect, 0.50-0.80 are a medium effect, and 0.20-0.50 are a small effect. Statistical analyses were conducted using SPSS V.18.

## RESULTS

A total of 48 nursing students participated in the study during May 2017 to July 2018. Twenty-four participants were assigned to group 1 and the other half were in group 2. A majority of participants (69%) were women and the median age was 25 years. Some subjects reported previous experience in basic airway management through didactic instruction in airway management or through practice with hands -on skill with mannequins. None of the participants reported having performed airway management with actual patients. There were no statistically significant differences between the two groups relative to previous airway knowledge or skill (Table 2).

**Table 2.** Baseline characteristics of the participants. (Group 1: oropharyngeal and nasopharyngeal airway skills by peer-to-peer with bag mask ventilation by standard instructor-led method, Group 2: standard instruction for oropharyngeal and nasopharyngeal airway by standard instructor-led method followed by bag mask ventilation instruction by peer-to-peer method)

|  |  | Total<br>[N 48, no(%)] | Group 1<br>[N 24, no(%)] | Group 2<br>[N 24, no(%)] | p-value |
| --- | --- | --- | --- | --- | --- |
| Age<br>(years old) | Mean (SD) | 24.88 (5.08) | 25.75 (4.12) | 25.04 (2.90) | 0.49 |
|  | Median<br>(min-max) | 25 (20-35) | 25 (20-34) | 25 (20-33) | 0.77 |
| Female |  | 33 (69.7) | 18 (75.0) | 15 (62.5) | 0.47 |
| Previous airway knowledge |  | 12 (25.0) | 8 (33.3) | 4 (16.7) | 0.22 |
| Previous airway skill |  | 36 (75.0) | 21 (87.5) | 15 (62.5) | 0.07 |
| Previous oropharyngeal airway skill |  | 22 (45.8) | 13 (54.2) | 9 (37.5) | 0.67 |
| Previous nasopharyngeal airway skill |  | 20 (41.7) | 13 (54.2) | 7 (29.2) | 0.84 |
| Previous bag mask ventilation skill |  | 32 (66.7) | 21 (87.5) | 11 (45.8) | 0.19 |

The scores of the participants who learned via the peer-to-peer method were compared to the standard teaching group scores. The upper limit of the 95% confidence interval (CI) for each skill was less than the established 10% margin (Table 3). These results demonstrate that for basic skill teaching, the peer-to-peer method is non-inferior to the standard one. With further analysis, it was clear that the peer-to-peer skill scores were higher than those of the students in the standard teaching group with a large calculated effect size for each skill. We calculated a Cohen’s *d* of 1.07 (p-value 0.002) for oropharyngeal airway, 1.14 (p-value <0.001) for nasopharyngeal airway and 0.81 (p-value 0.003) for bag mask ventilation.

**Table 3.** The procedural checklist scores separated by the learning method

| Procedural skill | Peer-to-peer<br>N=24,<br>[Mean (95% CI)] | Standard<br>N=24,<br>[Mean (95% CI)] | p-value | Effect size<br>(Cohen's *d*) |
| --- | --- | --- | --- | --- |
| Oropharyngeal airway<br>(maximum score = 12) | 10.42<br>(9.92-10.91) | 8.79<br>(8.00-9.58) | 0.002 | 1.07 |
| Nasopharyngeal airway<br>(maximum score = 12) | 11.54<br>(11.21-11.87) | 10.38<br>(9.58-10.9) | <0.001 | 1.14 |
| Bag mask ventilation<br>(maximum score = 16) | 14.92<br>(14.71-15.46) | 13.67<br>(12.69-14.31) | 0.003 | 0.81 |

The median knowledge score, from the 10-item MCQ, increased from 6 for pre-learning to 7 for post-learning (p-value 0.09). There was no significant difference between the pre-test and post-test scores between Group 1 and 2 (Table 4). There was one missing data point from a participant in Group II who didn’t complete the post-training survey. Therefore, the analysis was done on 47 participants for confidence, stress and perception level. For confidence level, participants in both groups reported higher confidence after learning in every airway skill (Figure 2). There was no statistically significant difference in post-learning confidence levels between the peer-to-peer and standard teaching groups with a p-value of 1.00 for oropharyngeal airway insertion, 0.77 for nasopharyngeal airway insertion, and 0.26 for bag mask ventilation. The stress during the learning sessions was significantly lower in the peer-to-peer group (stress score of 1.41 and 1.47 for peer-to-peer and standard instructor-led respectively, p-value 0.13). Further, the participants reported that the standard instructor-led method could increase their knowledge and skills better than the peer-to-peer group (Figure 3).

**Table 4.**
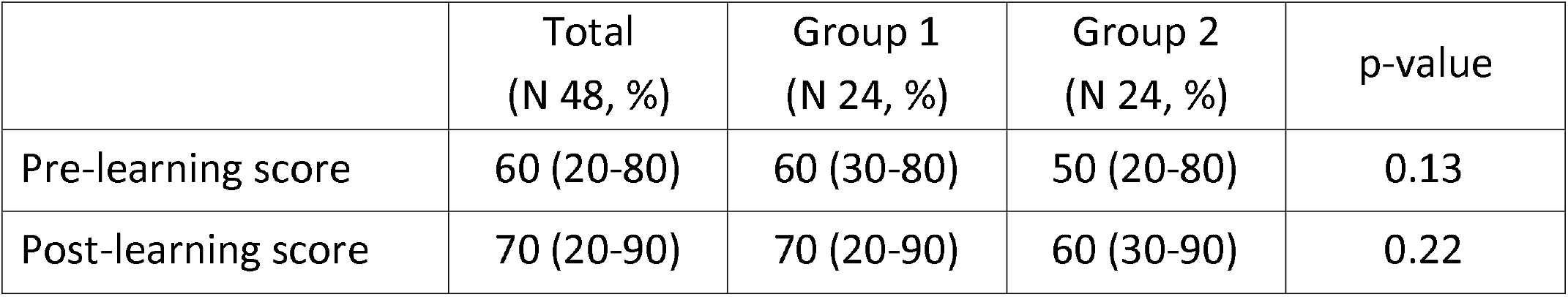
Median (minimum-maximum) of the knowledge percentage (10-item multiple choices question) (Group 1: oropharyngeal and nasopharyngeal airway skills by peer-to-peer with bag mask ventilation by standard instructor-led method, Group 2: standard instruction for oropharyngeal and nasopharyngeal airway by standard instructor-led method followed by bag mask ventilation instruction by peer-to-peer method)

**Figure 2.**
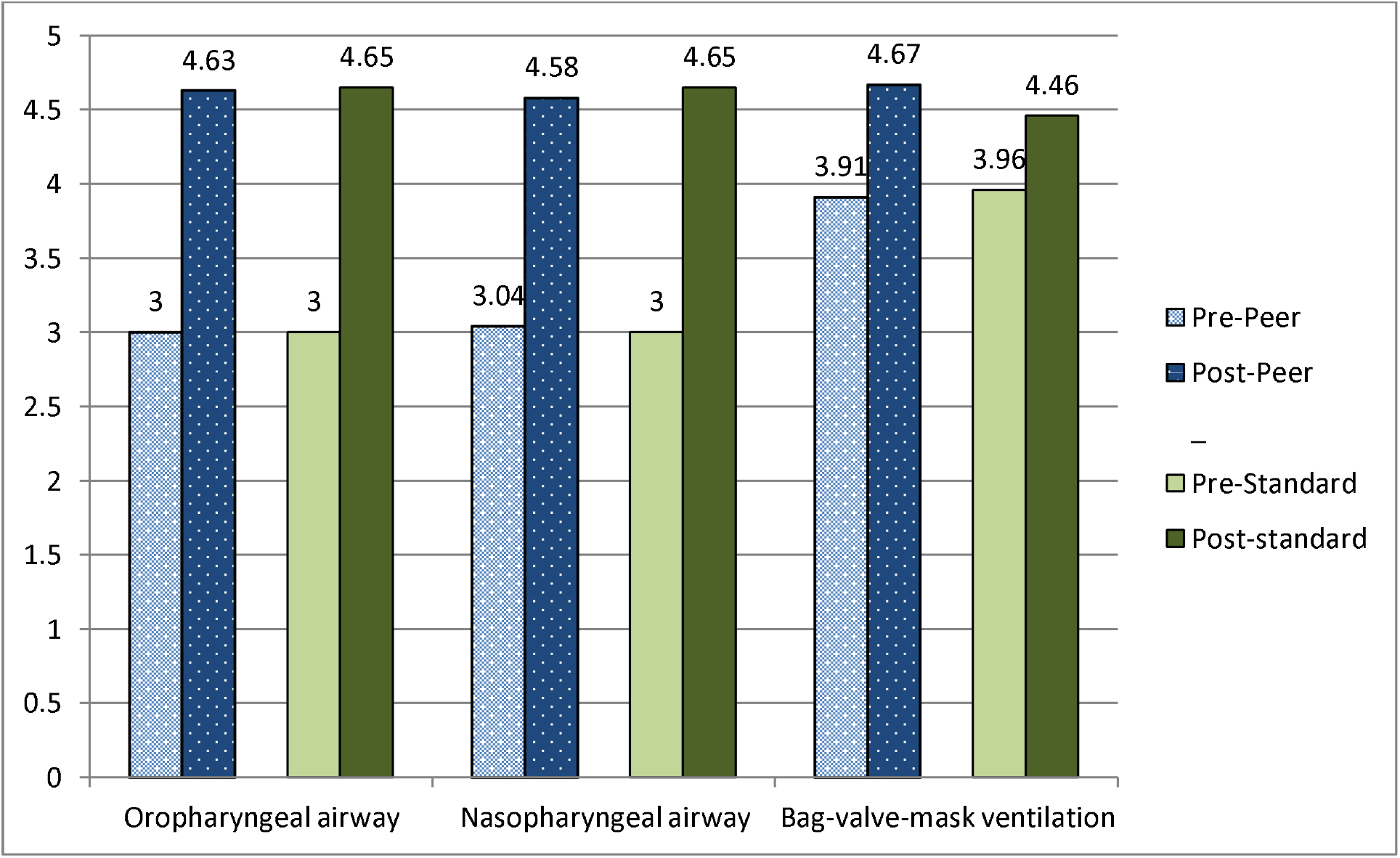
Confidence level before and after skill training separated by learning method (using Likert scale from 1 (strongly disagree) to 5 (strongly agree))

**Figure 3.**
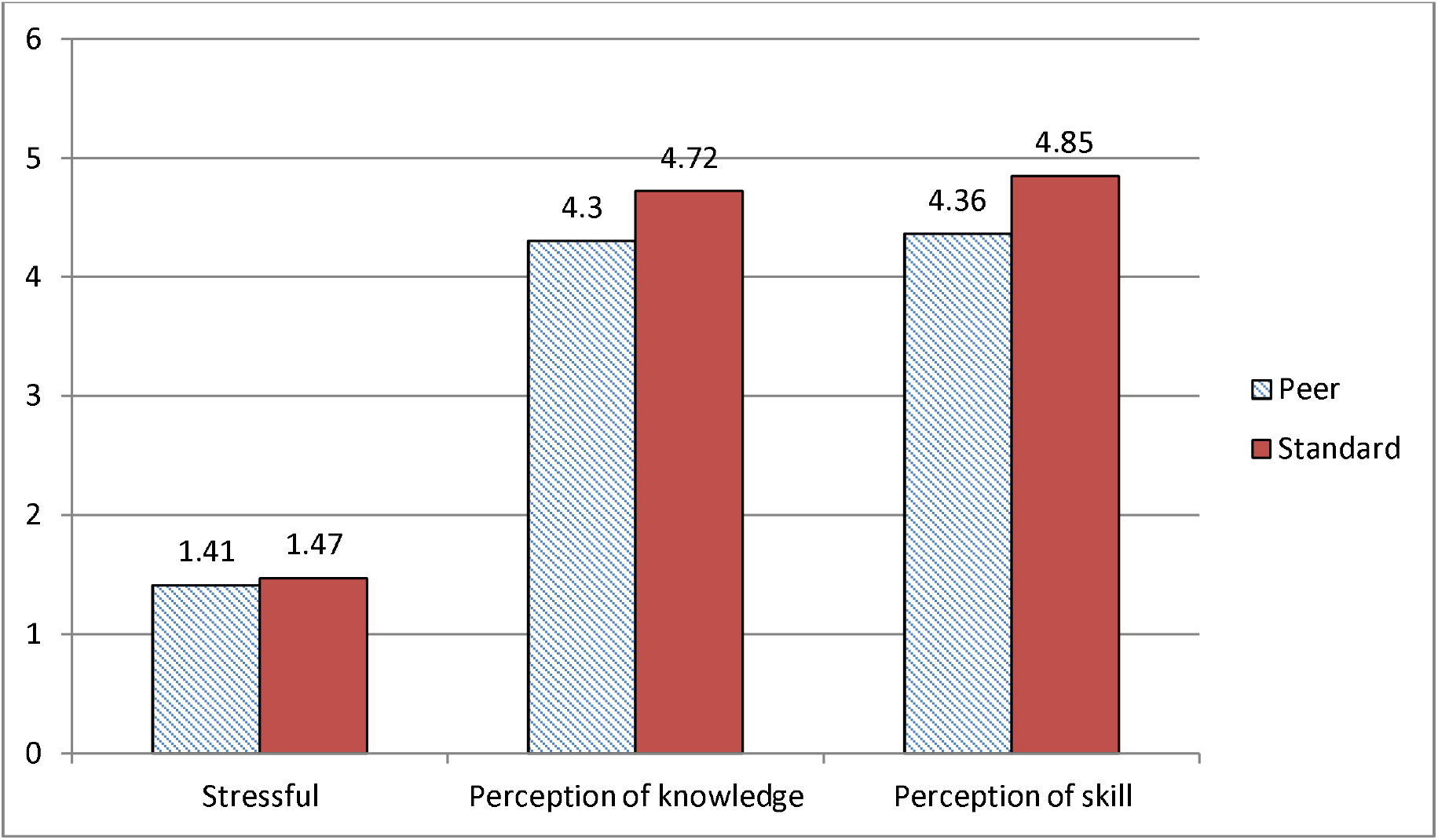
During class stress level and perception of knowledge and skill separated by learning method (using Likert scale from 1 (strongly disagree) to 5 (strongly agree))

After each learning session was completed, the participants were asked to choose their preferred learning method between standard instructor-led and peer-to-peer instruction. Thirty-two students (69.57%) chose standard instructor-led-teaching as the preferred method for basic airway management training. Among these students, 17 participants (53.12%) were from the group 1 assignment (peer-to-peer instruction first). The participants were also afforded the ability to add comments after their learning sessions (Table 5).

**Table 5.** Open comments of the participants about the learning method

| No | Comment |
| --- | --- |
| 1 | Less stressful with peer than “faculty” but both were equally informative |
| 2 | Glad I did this. |
| 3 | Teaching another peer is actually extremely helpful for learning yourself. I learned just as much by teaching my peer and practicing with my peer as I did when instructed by an expert. |
| 4 | I felt that peer learning was appropriate for the nasal/oral airway station. However I think it was less appropriate for BVM station where instructor critique of technique was more necessary. |
| 5 | Excellent |
| 6 | It was nice to have faculty around to be able to ask questions (ie related to real life/clinical situations). But I think that peer to peer teaching is an effective way of teaching a new skill. |
| 7 | For demonstration purposes, I believe both the instructor and peer to peer lead education was very similar. That being said, having an expert instructor is always better for more advanced questions being answered. |
| 8 | Both methods were helpful. |

## DISCUSSION

This study demonstrated the potential impact of an innovative teaching method on knowledge, skill (both airway and peer-to-peer instruction) and attitude (confidence). The teaching method combined self-instruction videos, peer-to-peer instruction, guided peer-feedback and debriefing with the use of the GAS model. This is a modification of what is sometimes described as a blended learning approach with the added innovation of the peer-to-peer instruction and a highly structured self-instruction video and feedback system.^8^ The skill performance score, knowledge score, and confidence level of the novice nursing students who learned basic airway management by the peer-to-peer method were not inferior to those who had learned by the standard instructor-led method. For the skill assessment score, the peer-to-peer group demonstrated significantly higher scores with a large effect size in every airway management skill category. The combination of the self-study video, peer-assisting learning, and structured debriefing system showed a significant benefit for skill performance. Self-instructed video teaching is effective in establishing psychomotor skills because of the ability to display a moving image.^9^ Learners tend to have a deeper understanding and longer-lasting memory with this teaching approach because video can arouse more than one sensory stimulation.^10^ Self-instruction videos have been demonstrated to be an effective alternative learning method for cardiopulmonary resuscitation and basic emergency skills for medical students and for baseline skill development for nursing students.^2,6,11^ The peer-assisted learning approach has also been demonstrated to improve under-graduate nurse participants’ outcomes in skill performance, knowledge level, and communication skills.^12,13^ Similar to findings in previous systematic reviews and meta-analyses, this study found that students who learned via peer-to-peer teaching methods had no difference in skill learning outcomes compared to those who were taught by standard instructional methods with expert instructors.^14^ Students also have an opportunity to develop their teaching skills in peer-to-peer instructional sessions which is a ‘value-added’ skill for their future practice. Therefore, the peer-to-peer teaching method should be considered as a valid alternative to standard instruction with the caveat that careful consideration must be taken in development of instructional materials as well as in quality monitoring. The structured GAS model debriefing method was added to the peer-teaching in order to enhance the student’s ability to give proper feedback. The GAS model is a simple and scalable debriefing method that can be rapidly learned by a novice for the purpose of stimulating peer reflection and debriefing of skill performance. The GAS model enhances learning by providing the student with an opportunity for self-discovery, self-reflection, and knowledge gap identification.^15^ Self-reflection has been shown to enhance self-assessment, self-monitoring, and skill performance.^16^

Peer-to-peer instruction methods were also shown to be non-inferior to standard instructor-led methods regarding knowledge and confidence levels. Both methods demonstrated higher knowledge and confidence scores on post-knowledge tests. Approximately 70% of the participants indicated that the standard-teaching method was preferred despite our finding that skill attainment was greater after peer-to-peer instruction. Participants reported several advantages to peer-to-peer teaching including less stress and the opportunity to learn through teaching their peer.

The standard instructor-led teaching method has some disadvantages. These include the need for high student to faculty ratios (in many institutions), the high cost of instructors and the potential for lack of standardization among instructors. Based on previous online surveys of 833 clinical teachers, the most common challenge for instructors in providing high quality teaching is lack of adequate preparation time.^17^ Thus an added disadvantage is the need for additional and sometimes extensive time committed to instructor preparation. Self-instruction videos and peer-to-peer instruction combined with a guided peer-to-peer structured debriefing feedback model demonstrated significant benefit in teaching basic airway management skills in novice students with minimal preparation time. It also had the significant collateral benefit of reduced cost and decreased instructor workload.

## Limitations

This study demonstrated a non-inferiority effect for the immediate post peer-to-peer teaching period. However, the long-term knowledge and skill retention of the participants was not tested. Although this teaching method appears to be highly influential in the retention of knowledge and skills for students in the near term; it has not been tested whether peer-to-peer teaching is non-inferior to standard expert teaching in maintaining long-term knowledge and skills. A future study should include and examine this outcome.

Although the peer-to-peer method proved to be effective in teaching basic airway management for novice nursing students, the ability to generalize the method to other skills and participants was not analyzed. It is important to consider that peer-to-peer teaching may only be effective with a certain level of skills and participants. It may not be appropriate to use peer-to-peer teaching for more complex knowledge and skills. It is also recommended that an in-depth analysis and ongoing evaluation of the peer-to-peer teaching course be performed to ensure that the quality of instruction is as intended. Further validation of the self-instruction video is also imperative as it is the main contributor to the standardization of education.

A last limitation is the established culture of the ‘expert instructor’ being the gold standard for educational effectiveness. This is reflected in student perceptions reported in this study that the instructor led sessions were preferred and in the open narrative comments which indicated this bias (Table 5). It may be necessary to include evidence-based findings on peer-to-peer instruction in pre-course materials in order to help learners understand that this is a cultural bias and not based in fact.

## CONCLUSION

In conclusion, this study demonstrated that novice nursing students who learned basic airway management skills with the combination of self-instruction videos, peer-to-peer instruction, guided peer-to-peer feedback and structured debriefing using the GAS model was not inferior to standard instructional methods in the domains of knowledge, skill and attitude (confidence). Futher analysis revealed higher and statistically significant performance scores for the airway management skills of oral airway insertion, nasal airway insertion and bag-mask ventilation with large effect sizes for peer-to-peer instruction versus standard instructor-led methods.

## Data Availability

All data relevant to the study are included in the article or uploaded as supplementary information

## Contributors

The Peter M. Winter Institute for Simulation, Education and Research (WISER) provided the simulation resources for the learning sessions in the study.

## Ethics approval

IRB# PRO17040484, Exempt approved under the University of Pittsburgh.

## Provenance and peer review

Not commissioned; externally peer reviewed.

## Data sharing statement

No additional unpublished data.

